# The relation between COVID-19 vaccinations and public governance to improve preparedness of next pandemic impacts and crisis management: a global study

**DOI:** 10.1101/2022.04.10.22273663

**Authors:** Mario Coccia, Igor Benati

**Affiliations:** CNR -- NATIONAL RESEARCH COUNCIL OF ITALY; Collegio Carlo Alberto, Via Real Collegio, 30-10024 Moncalieri (Torino, Italy)

**Keywords:** COVID-19, Coronavirus infections, SARS-CoV-2, Public governance, Government effectiveness, Regulatory quality, Rule of law, Health systems, Health governance, Pandemic prevention, Crisis management

## Abstract

The goal of this study is to analyze the relationship between COVID-19 vaccinations and public governance performing a global analysis of more than 110 countries worldwide. Methodology applies the Independent Samples *T*-Test that compares the means of two independent groups (countries with high/low level of vaccinations) to determine whether there is statistical evidence that the associated population means of indicators of public governance are significantly different. Findings suggest that high levels of governance can support a better function of health systems in the rollout of vaccinations to cope with COVID-19 pandemic crisis. This study may assist long-run policy of governments to improve good governance and health systems of countries in order to reinforce the preparedness to face next pandemic threats and in general future crisis management in society.

## Introduction

Coronavirus disease 2019 (COVID-19) is an infectious disease caused by the novel Severe Acute Respiratory Syndrome Coronavirus 2 (SARS-CoV-2), which appeared in late 2019 (Coccia, 2020, 2020a, 2020b, 2020c). COVID-19 is still circulating in 2021 with new variants (e.g., Delta) of the novel influenza coronavirus that continue to be a constant pandemic threat in various countries worldwide (Johns Hopkins Center for System Science and Engineering, 2021). The alarming levels of spread and severity of COVID-19 have supported the development of many vaccines in 2020 (World Health Organization, 2021). Mass vaccination is one of the policy responses to cope with the COVID 19 pandemic (Abbasi, 2020; DeRoo et al., 2020; Dooling et al., 2020; Frederiksen et al., 2020; Harrison and Wu, 2020). In December 2020, large vaccination campaigns started in many countries around the world (Freed, 2021). In some countries, the vaccination campaign was rapidly successful, resulting in a high number of vaccine doses administered in a short time (Cylus et al., 2021). In other countries, the rollout of vaccinations has been slower. The explanation of these differences is linked to manifold economic, socio-cultural and political-administrative factors (Bontempi et al., 2021; Bontempi and Coccia, 2021; DeRoo et al., 2020; Durmus, 2021). Economic factors are basic since mass vaccination campaign involves enormous public investment to be organized (Ethgen et al., 2019; Faes et al., 2020). The wealth of nations can be a discriminating factor for success of health systems and health policy responses (Durmus, 2021). Studies suggest that higher-income countries perform better than low-income ones in vaccinating their citizens (Ataguba et al. 2016). Vaccination rates are also influenced by socio-cultural factors, such as house living conditions (cf., Kusuma et al. 2010; Mitchell et al. 2009), education, religious beliefs (Soura et al. 2013), gender-based inequity (Pande, 2003), information and communication (Logan et al. 2018; Su, 2021). In this context, political and administrative factors are relevant aspects for crisis management of pandemic threats, but they are poorly investigated (Glatman-Freedman and Nichols, 2012; Coccia, 2021, 2021d, 2022; Kluge et al., 2020). Some studies show that public governance is a relevant factor to build policy responses against COVID-19 pandemic (Cylus et al., 2021; Kluge et al., 2020; Sagan, 2021). Janssen and van der Voort (2020) highlighted the role of an agile and adaptive governance in crisis response in Dutch experience. Zhang and Zhang (2020) also stress the importance of public governance to combat the pandemic. Anttiroiko (2021) analyzes how socio-economic context, institutional arrangements, culture, and technology level can affect national responses to the pandemic of Eastern and Western countries (Ardito et al., 2021). The study reveals that Asian countries reflect proactivity, whereas Western countries provide reactive policy responses. In general, crisis management of COVID-19 pandemic is based on effective multi-level governance, combining both national and urban strategies to improve safety in society (Abuza, 2020; Anttiroiko, 2021). However, there is little evidence of how public governance can be associated with the success of a vaccination campaign. This study aims to propose a global analysis for a broader and more argued demonstration of the relation between good governance and effectiveness of the vaccination campaign of COVID-19 between countries.

### Theoretical framework

The term governance, which denotes the action of ‘giving direction’, has experienced great fortune in public administration literature in the last 20 years (Rhodes, 1996; Pierre, 2000; Bevir, 2010; Ansell and Torfing, 2016). In general terms, governance refers to country capacity to build structure and process for steering and coordinating policymaking, identifying some effective means of deciding upon collective goals and then finding the means of reaching those goals (Guy Peters, 2014). From a functional point of view, governance can be defined as a government’s ability to make and enforce rules and to deliver services, regardless of whether that government is democratic or not (Fukuyama, 2013). In a broad sense, governance is about the culture and institutional environment in which citizens and stakeholders interact among themselves and participate in public affairs (International Bureau of Education, 2021; Coccia, 2018). Worldwide Governance Indicators (2021) state that governance consists of the traditions and institutions by which authority in a country is exercised. In particular, the public governance includes the capacity of the government to effectively formulate and implement sound policies; and the respect of citizens and the state for the institutions that govern economic and social interactions among them.” In society, this specific function is shared between government, public administration and economic subjects. Kaufmann et al. (2009) synthetically defined governance as: ‘traditions and institutions by which authority in a country is exercised’. This definition was operationalized by the World Bank in the system of Worldwide Governance Indicators (WGI), given by six aggregate indicators: (1) voice and accountability (‘the extent to which a country’s citizens are able to participate in selecting their government, as well as freedom of expression, freedom of association, and free media’); (2) political stability and the absence of violence; (3) government effectiveness; (4) regulatory quality; (5) the rule of law; and (6) control of corruption (World Bank, 2007, 2007a). Good governance is the combination of transparent and accountable institutions, strong skills and competence, and a fundamental willingness to do the right things; all aspects that enable a government to deliver services to its people efficiently (Addink, 2019; Coccia, 2018; Hood and Guy Peters, 2003; World Bank, 2007, 2007a). Good governance has a whole range of positive effects on economic growth (Kaufmann et al., 1999; Coccia, 2005, 2017, 2018b; Coccia and Bellitto, 2018), enhancing public confidence in the institutions and improving public services (Jameel et al., 2019). Glatman et al. (2010) highlight the role of country-level governance in the context of new vaccine introduction to poor African countries. The study shows that poor African counties with higher levels of governance scores were more likely to administer new vaccines, suggesting the importance of governance factors in the implementation of vaccination programs (cf., Glatman-Freedman and Nichols, 2012). In addition, superior country-level governance was formerly demonstrated to have a substantial impact on health-related investments in developing countries (Coccia, 2021). In fact, country-level governance was found to be a stronger predictor of the initial introduction of new vaccines to poor African nations than healthcare-related financial indicators (Glatman et al. 2010; Glatman-Freedman and Nichols, 2012). The ability to organize a vaccination campaign to cope with COVID-19 pandemic crisis can be compared to public service organization in a short period of time in a context of crisis management (Coccia, 2021).In general, country-level governance may also affect the infrastructure required for the successful rollout of vaccine programs, such as the ability to reach distant locations, cold chain capacity, safe disposal of used syringes and needles, and adequate numbers of trained personnel (Freed, 2021). In this context, next section presents the study design to analyze the relationship between COVID-19 vaccinations and public governance performing a global analysis of more than 110 countries to provide preliminary lessons learned to cope with pandemic threats.

## Materials and methods

### 1.1 Research setting and sample

The study is a global analysis of *N*=112 countries worldwide. Period under study is March 2021, during the initial phase of mass vaccination between countries because the goal is to determine factors of public governance associated with countries having a faster reaction capacity to rollout vaccination for a vast population in a short period of time to support effective and timely policy responses for combatting the novel coronavirus and constraining negative effects of COVID-19 pandemic crisis and future pandemics of similar infectious diseases.

### 1.2 Measures and sources

□ Doses of vaccines administered × 100 inhabitants at February-March 2021. Doses of vaccines refer to the total number of vaccine doses, considering that an additional dose may be obtained from each vial (e.g., six doses for Pfizer BioNTech® Comirnaty), whereas number of doses administered refers to any individual receiving any dose of the vaccine (cf., Freed et al., 2021; Oliver et al., 2020). Source: Our World in Data (2021).
□ Number of COVID-19 infected individuals (%) is measured with confirmed cases of COVID-19 divided by population of countries under study on 14 December 2020. Source of data: Johns Hopkins Center for System Science and Engineering (2021).
□ Number of COVID-19 deaths is measured with fatality rate (%) of COVID-19 given by deaths divided by total infected individuals in countries on 14 December 2020. Source of data: Johns Hopkins Center for System Science and Engineering (2021).

This study uses indicators of public governance of countries by Worldwide Governance Indicators (2021) over the year 2019, given by: Government effectiveness, Regulatory quality and Rule of law. In particular,

□ Government effectiveness in 2019: Government effectiveness captures perceptions of the quality of public services, the quality of the civil service and the degree of its independence from political pressures, the quality of policy formulation and implementation, and the credibility of the government’s commitment to such policies (Worldwide Governance Indicators, 2021).
□ Regulatory quality 2019: Regulatory quality captures perceptions of the ability of the government to formulate and implement sound policies and regulations that permit and promote private sector development (Worldwide Governance Indicators, 2021).
□ Rule of law 2019: Rule of law captures perceptions of the extent to which agents have confidence in and abide by the rules of society, and in particular the quality of contract enforcement, property rights, the police, and the courts, as well as the likelihood of crime and violence (Worldwide Governance Indicators, 2021).

### 1.3 Data analysis procedure

*Firstly*, data are analyzed with descriptive statistics of variables given by arithmetic mean (M) and standard deviation (SD). In addition, the normality of the distribution of variables to apply correctly parametric analyses is analyzed with skewness and kurtosis coefficients; in the presence of not normal distributions, variables are transformed in logarithmic scale to have normality and apply correctly parametric analyses. The arithmetic mean of doses of vaccines administered × 100 inhabitants in March 2021 of all sample *N*=112 countries, given by 9.93, is used to divide the sample in two sub-samples for a comparative analysis (Coccia and Benati, 2018; Coccia, 2018a):

□ group 1: countries with a high level of vaccine doses administered >9.93 per 100 inhabitants, *N*=42

□ group 2: countries with a low level of vaccine doses administered ≤9.93 per 100 inhabitants, *N*=70

*Secondly*, the difference of arithmetic mean of variables between group 1 and 2 is analyzed with the Independent Samples t-Test. In particular, the Independent Samples t-Test compares the means of two independent groups in order to determine whether there is statistical evidence that the associated population means are significantly different. The assumption of homogeneity of variance in the Independent Samples *T* Test --i.e., both groups have the same variance -- is verified with Levene’s Test based on following hypotheses:

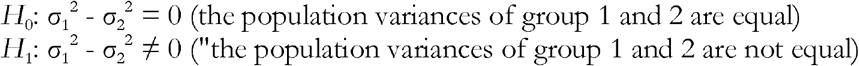

The rejection of the null hypothesis in Levene’s Test suggests that variances of the two groups are not equal: i.e., the assumption of homogeneity of variances is violated. If Levene’s test indicates that the variances are equal between the two groups (i.e., *p*-value large), equal variances assumed. If Levene’s test indicates that the variances are not equal between the two groups (i.e., *p*-value small), the assumption is that equal variances are not assumed. After that, null hypothesis (*H*’_0_) and alternative hypothesis (*H*’_1_) of the Independent Samples *t*-Test are: *H*’_0_: µ_1_ = µ_2_, the two-population means are equal in countries with a higher and lower GDP per capita *H*’_1_: µ_1_ ≠ µ_2_, the two-population means are not equal in countries having a higher and lower GDP per capita *Thirdly*, bivariate Pearson correlations were used to verify relationships between variables, with the degree of association determined by the coefficient of correlation. One-tailed tests of significance for correlation were computed to consider the associations between variables in the last year available in the databases. Finally, it is analyzed the relation of the dependent variable (Doses of vaccines administered × 100 inhabitants in 2021) on explanatory variables of government effectiveness, regulatory quality and rule of law. The specification of the model of simple regression is given by:

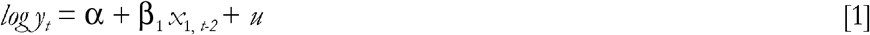

where

*y* = dependent variable (Doses of vaccines administered × 100 inhabitants in 2021)

*x* _1_ *=* explanatory variables of government effectiveness, regulatory quality and rule of law 2019

α = constant; β = coefficient of regression; *u* = error term

Statistical analyses are performed with the Statistics Software SPSS® version 26.

## Results

Firstly, descriptive statistics of variables shows that Doses of vaccines and fatality rates have not normal distribution and for this reason these variables have a log transformation to apply appropriate parametric analyses.

Table 2 shows that countries with high activity of vaccine doses administered (average of bout 3 per 100 people) in general have also better indicators of governance (government effectiveness, regulatory quality and rule of law) than countries with a low activity of vaccine doses administered (average value of 2.4 doses per 100 inhabitants).

**Table 1.** Descriptive statistics, N=112 countries

| | Doses of vaccines<br>administered ×<br>100 inhabitants<br>March 2021 | Confirmed<br>cases %<br>population<br>March 2021 | Fatality<br>rates March<br>2021 | Government<br>effectiveness<br>2019 | Regulatory<br>quality<br>2019 | Rule of law<br>2019 |
| --- | --- | --- | --- | --- | --- | --- |
| Mean | 9.93 | 0.03 | 0.02 | 0.37 | 0.36 | 0.28 |
| Std. Dev. | 16.23 | 0.03 | 0.01 | 0.84 | 0.89 | 0.93 |
| Skewness | 3.80 | 0.95 | 1.85 | 0.28 | -0.04 | 0.22 |
| Kurtosis | 18.02 | 0.72 | 6.89 | -0.67 | -0.31 | -0.61 |

**Table 2.** Descriptive statistics considering groups with high/low level of doses of vaccines administered

|  | HIGH level of vaccine doses<br>adm. > mean 9.93,<br>N= 42 |  | LOW level of vaccine doses<br>adm. < mean 9.93,<br>N=70 |  |
| --- | --- | --- | --- | --- |
|  | Mean | Std. Dev. | Mean | Std. Dev. |
| Doses of vaccines administered × 100<br>inhabitants, March 2021 | 22.46 | 21.04 | 2.41 | 2.82 |
| Confirmed cases % Population, March 2021 | 0.05 | 0.03 | 0.02 | 0.03 |
| Fatality rates, March 2021 | 0.02 | 0.01 | 0.02 | 0.01 |
| Government effectiveness, 2019 | 0.97 | 0.68 | 0.00 | 0.71 |
| Regulatory quality, 2019 | 1.02 | 0.64 | -0.03 | 0.77 |
| Rule of law, 2019 | 0.99 | 0.69 | -0.14 | 0.79 |

The difference of arithmetic mean of between countries with a high/low activity of vaccine doses administered is analyzed with Independent Samples *T* Test (Table 3). In this context, results show that there is a significant difference in average values of variable under studies between the two groups of countries having a high/low activity of vaccine doses administered, supporting the results described in table 2: i.e., a higher efficiency to administer doses of vaccines (and health system) is associated with a better general governance of countries.

**Table 3.**
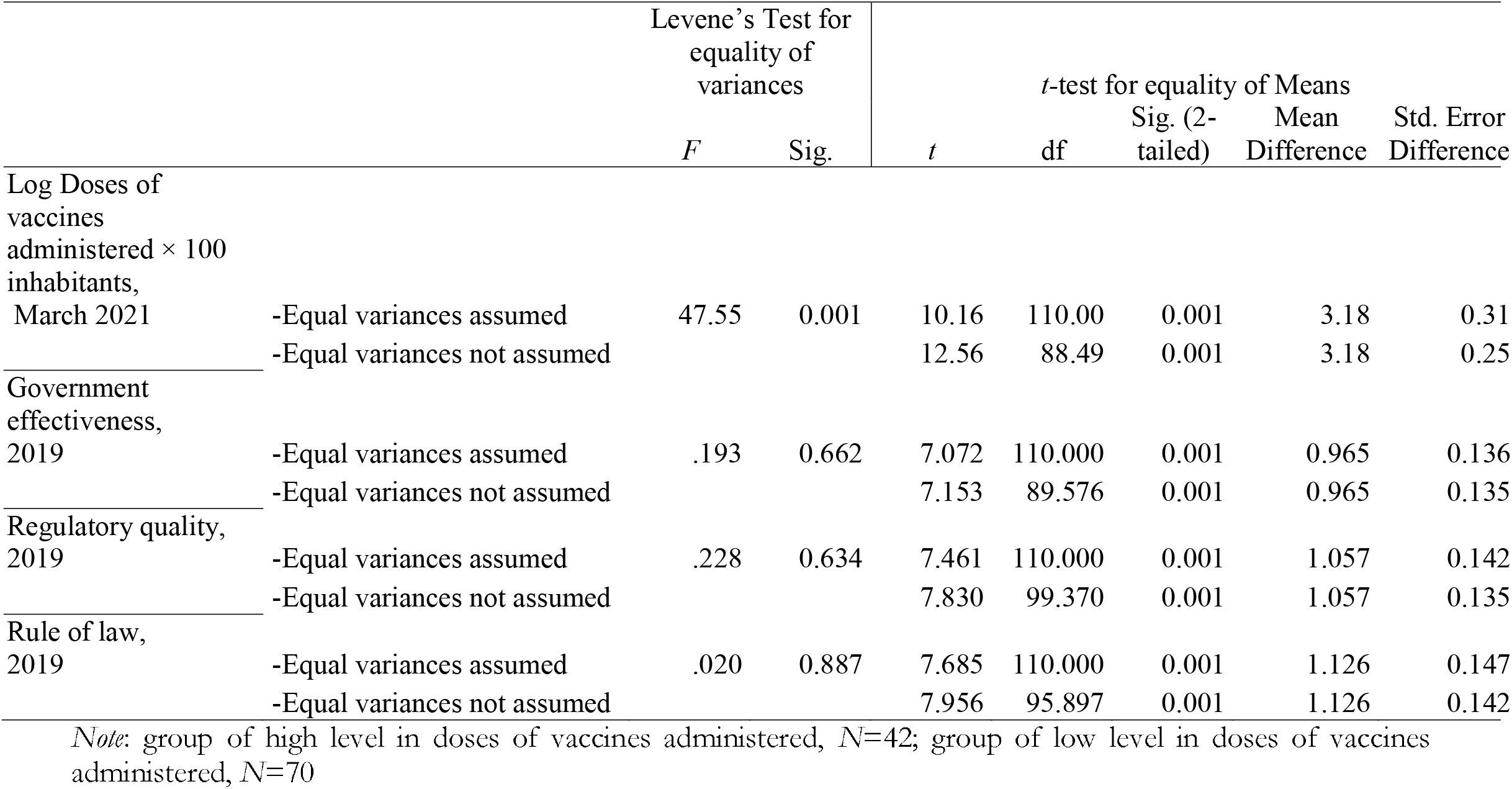
Independent Samples Test between groups with high/low level of doses of vaccines administered

Table 4 shows bivariate correlation of variables under study: Doses of vaccines administered × 100 inhabitants March 2021 have a high positive association with Government Effectiveness (*r*=.56, *p*-value <.01), regulatory quality (*r*=.58, *p*-value <.01), and rule of law (*r*=.55, *p*-value <.01).

**Table 4.** Correlation

| | Log<br>Doses of vaccines<br>administered × 100<br>inhabitants<br>March 2021 | Government<br>effectiveness<br>2019 | Regulatory<br>quality<br>2019 | Rule of law<br>2019 |
| --- | --- | --- | --- | --- |
| Log Doses of vaccines<br>administered × 100 inhabitants,<br>March 2021 | 1 | .555** | .580** | .556** |
| Government effectiveness, 2019 |  | 1 | .939** | .952** |
| Regulatory quality, 2019 |  |  | 1 | .938** |
| Rule of law, 2019 |  |  |  | 1 |

Note: \*\* Correlation is significant at the 0.01 level (1-tailed).

Finally, table 5 shows the estimation of parameters of loglinear models between dependent variables (based on Log Doses of vaccines administered × 100 inhabitants March) and explanatory variables of governance (see, Appendix for interpretation of the coefficients of regression here). The coefficient of regression in model indicates that, an increase of 1 unit of government effectiveness, it improves the expected administration of vaccines by approximately 3.34 units (*p-value* = .001), whereas an increase of 1 unit of regulatory quality, it increases the expected giving of doses of vaccines by 3.27 units, finally an increase of 1 unit of the rule of law in countries, it increases the expected giving of doses administered of vaccines against COVID-19 by approximately 2.79 (*p-value* = .001). The coefficient R^2^ indicates that about 30% of the variation of administration of doses of vaccines can be attributed (linearly) to indicators of governance under study here (cf., Figure 1).

**Table 5.** Regression analyses

| *Dependent variable*<br>*Log Doses of vaccines administered*<br>*× 100 inhabitants, March 2021* | *Explanatory variables* |  |  |
| --- | --- | --- | --- |
|  | Government effectiveness<br>2019 | Regulatory quality<br>2019 | Rule of law<br>2019 |
| Constant $\alpha$<br>(St. Err.) | .357***<br>(.191) | .366*<br>(.186) | .517**<br>(.183) |
| Coefficient $\beta$<br>(St. Err.) | 1.467<br>(.210) | 1.451***<br>(.195) | 1.333***<br>(.190) |
| $R^2$<br>(St. Err. of Estimate) | .301<br>(1.857) | .330<br>(1.818) | .303<br>(1.854) |
| *F* | 48.84*** | 55.649*** | 49.216*** |
| *N* | 112 | 112 | 112 |

Significance: \*\*\* $p < 0.001$ ; \*\* $p < 0.01$ ; \* $p < 0.05$

**Figure 1.**
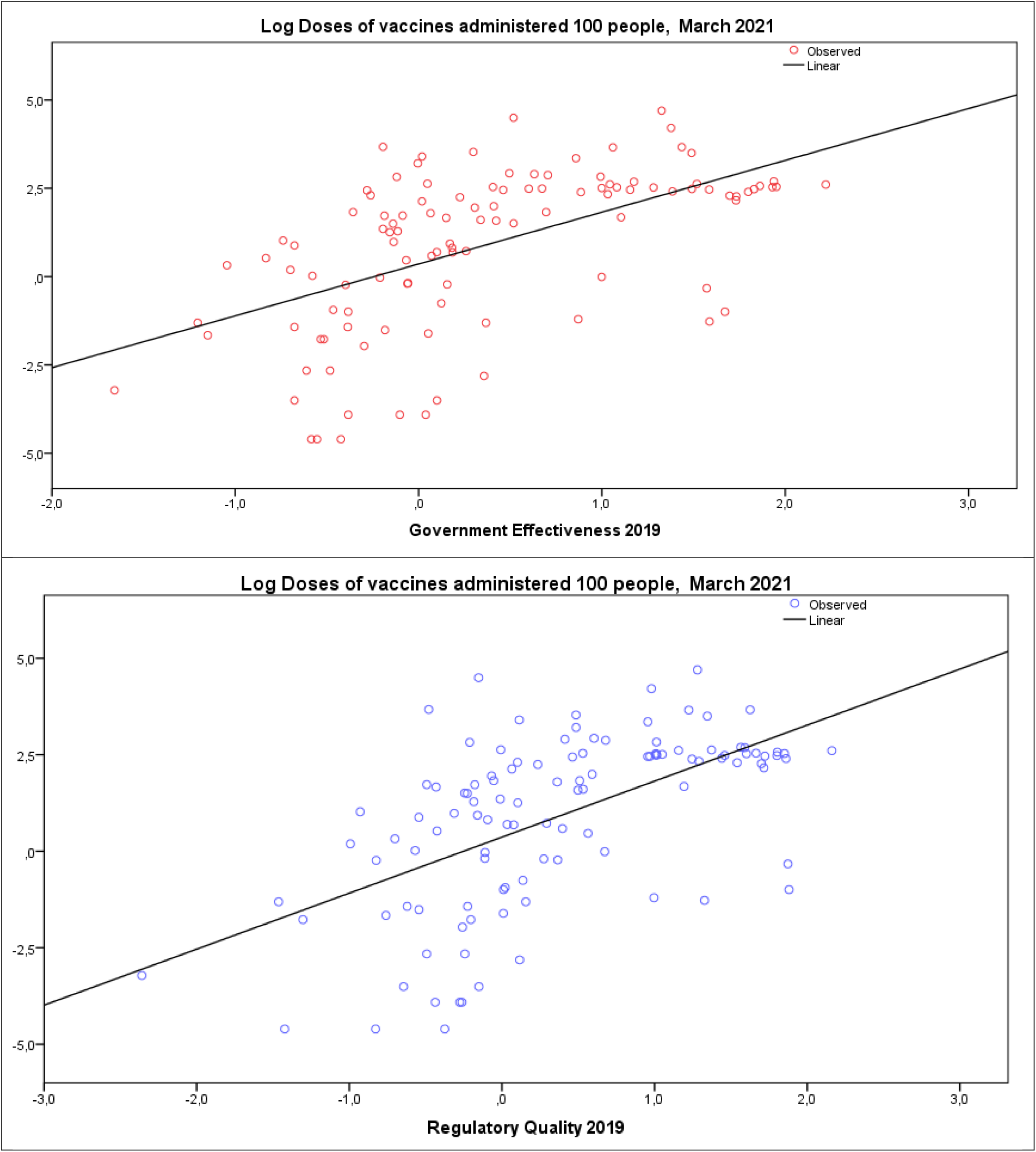
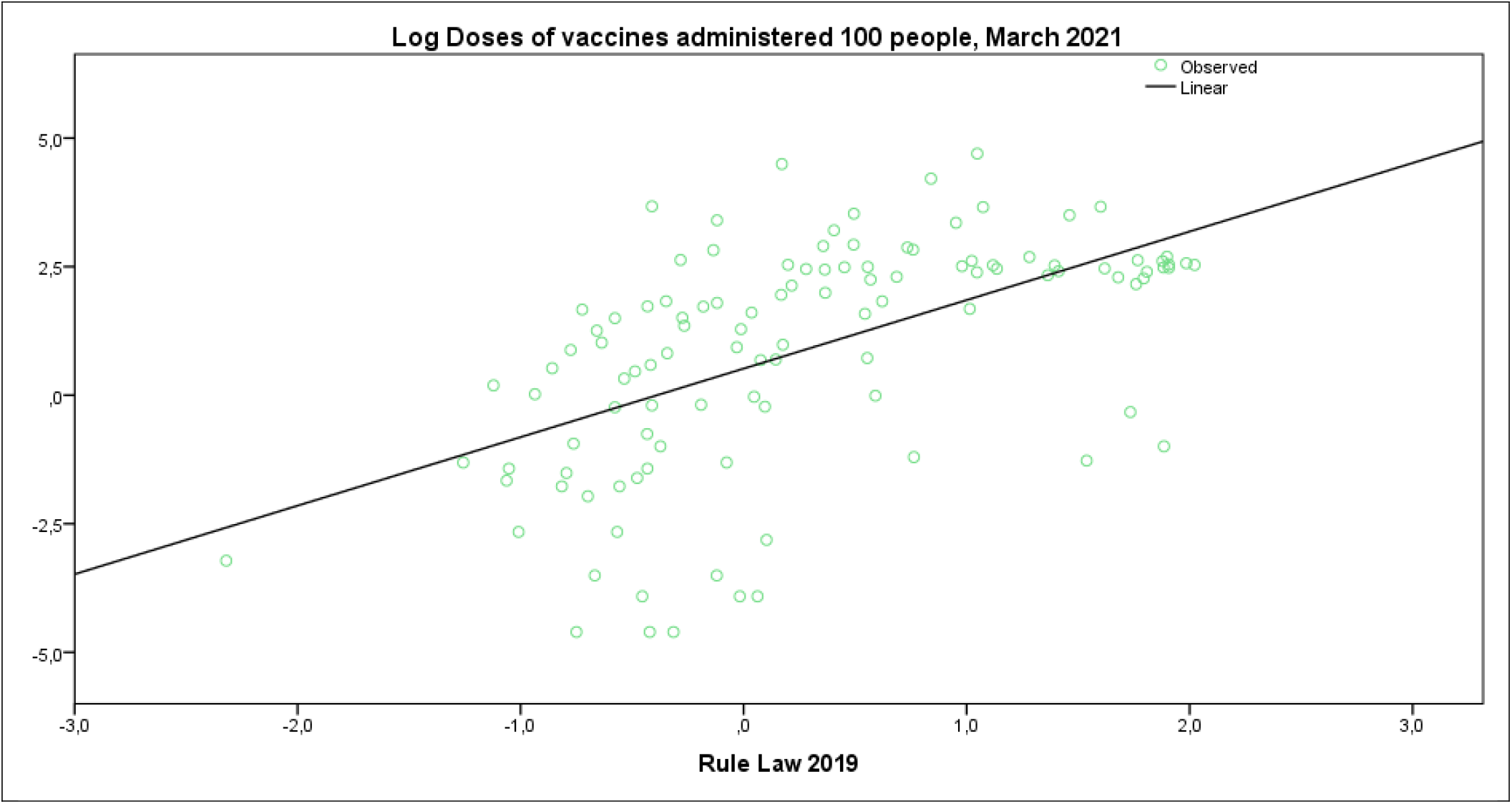
Regression lines of doses of vaccines administered on Government effectiveness, Regulatory quality and Rule of law 2019

## Discussion

This study shows how the different public governance capabilities of countries can justify their different performances in the management of vaccination campaigns to face the novel coronavirus and constraining negative effects of COVID-19 pandemic crisis and future pandemics of similar infectious diseases. The major findings here are that countries with higher values in main governance indicators (government effectiveness, regulatory quality and rule of law), achieve a higher vaccination rate of the population at the initial stage of the campaign.

Firstly, results here support the idea that good governance has a positive effect on the delivery of public services to citizens (Patton, 2008). In fact, if we consider COVID 19 vaccination campaign as service provided to the population of a country, which must be conducted in the most effective way and in the shortest possible time, our results show that countries which have higher vaccination rates, have also have higher values of all governance indicators under study. These results indicate a greater capacity of countries having a good governance to organize services and reach out to their citizens, motivating them to participate in vaccination efforts (Ali and Altaf, 2021; Cylus et al., 2021; Freed, 2021). Our results also highlight how rule of law and regulatory quality play a role in determining the effectiveness of the delivery of public services to citizens.

Secondly, the results of our study support the theory that good governance is a feature of resilient countries (Briguglio et al., 2009; Briguglio, 2016). These results here endeavor to detect governance factors in countries that develop resilience for increasing efficiency of crisis management in the short run to support mass vaccinations in the presence of pandemic threat. The concept of a resilient recovery underpins many national and international recovery plans (Sagan et al., 2020). Williams et al. (2020) argue that effective responses to public health emergencies should rely on translating rapidly emerging research into timely, evidence-informed policy and practice. Resilient systems to pandemic shocks must have strong governance structures driven by adequate and effective leadership that engages with the communities, listens and adjusts to population needs. Good governance can support health system preparedness in the presence of pandemic and environmental threats (Sagan et al., 2020; Kluge et al., 2020). Moreover, countries with constant investment in health sector can reduce mortality and morbidity among the population in the presence of pandemic threats (Coccia, 2021, Kluge et al., 2020). Sagan et al. (2020) also argue that effective governance is a critical factor to a resilient response in the presence of crisis among health system functions. In fact, critical aspects of resilient responses of countries to COVID-19 pandemic are two: 1) appropriate and effective governance and 2) technical capacity to respond in a short period of time. In particular, governance is increasingly a necessary condition for any effective response to cope with pandemic responses, such as lockdown (cf., Coccia, 2021a, 2021b, 2021e, 2022a, 2022b, 2022c). Sagan et al. (2020) consider a broad concept of governance that is not limited to health system alone. In fact, governance is a complex system of elements rather than a standalone function and creates the background to support other functions of nation and its government to work properly and strengthen health, economic and social systems in the presence of pandemic threats. As a consequence, public policies directed to enhance resilience of health systems have to be based on better governance of countries for supporting both policy responses of short run and long run interventions to prevent future social and health issues.

## Conclusions and prospects

COVID-19 and future epidemics of novel influenza viruses pose, increasingly, a serious threat to national security and public health (Coccia, 2022d). An influenza pandemic can occur at any time with little warning and any delay in detecting a novel influenza strain, sharing of influenza virus samples and in developing, producing, distributing, or administering vaccines could result in significant additional morbidity and mortality and deterioration of socio-economic systems. The global response to COVID-19 pandemic has pushed the boundaries on what is possible for rapid pandemic response in several areas, including healthcare system, vaccine research, development, manufacturing, distribution, allocation, and administration. To adequately prepare for, prevent, detect, and respond to both epidemics and inevitable pandemics, it is important to reinforce the governance of countries supporting domestically based seasonal and pandemic preparedness efforts also by collaborating with domestic and international stakeholders (Coccia, 2021b, 2021c). New strategies of nations in the presence of environmental threats have to be highly responsive, flexible, resilient, scalable, and more effective for reducing the impact of seasonal and pandemic influenza viruses. Execution of this strategic approach over the next ten years will require a better governance, innovative partnerships, financial investments, and efficient utilization of resources (U.S. Department of Health and Human Services, 2021; Jeyanathan et al., 2020). In short, policies having agility and speed of responses has to be based on a better governance of countries that is a vital factor for crisis management to cope with epidemic/pandemic threats (Chang et al., 2020; Janssen and van der Voort, 2020; Renardy et al., 2020). In the presence of a good governance, Evans and Bahrami (2020) pinpoint that super-flexibility can be an appropriate approach to cope with COVID-19 pandemic in which decision making is oriented to versatility, agility, and resilience.

Overall, then, suggested results of improving governance of countries here can help policymakers to reinforce institutions to cope with infectious diseases and to prevent future outbreaks of the COVID-19 and other new viral agents in future (Coccia, 2019). However, the proposed results here have the limit to consider some indicators of governance, but other confounding factors should be included in future development of this study. Therefore, to conclude, this study encourages further investigations for developing comprehensive analyses directed to consider institutional and socio-economic factors, and not only parameters related to medicine, to support policy resp9onses and design of short-run and long-run strategies to prevent future epidemics and/or to contain the negative impact of infectious diseases on public health, economy and society.

## Data Availability

All data produced in the present study are available upon reasonable request to the authors and online

## Appendix

Interpreting coefficients of regression in loglinear models.

Note that in loglinear model, *exp*(β)=1+ β, hence β=exp(β) − 1;

As a consequence, coefficients of regression in table 5 indicate:

| Estimated coefficient of regression | Government effectiveness | Regulatory quality | Rule of law |
| --- | --- | --- | --- |
| b | 1.47 | 1.45 | 1.33 |
| $\exp(b)$ | 4.34 | 4.27 | 3.79 |
| $\exp(b)-1$ | 3.34 | 3.27 | 2.79 |

